# Socially Grounded Exemplars Improve Synthetic Conversations for Health-Related Social Needs Navigation

**DOI:** 10.64898/2026.01.30.26345239

**Authors:** Syed-Amad Hussain, Daniel I. Jackson, Samanvith Thotapalli, Marissa B. McClellan, Madeleine Stanco, Grace Varney, Sterling Gleeson, Florencia Nugroho, William Leever, Eric Fosler-Lussier, Emre Sezgin

## Abstract

Health-Related Social Needs (HRSNs) significantly impact health outcomes, yet traditional care often fails to address them effectively. While conversational agents offer scalable support, their deployment is hindered by privacy risks and a lack of specialized training data for clinical applications. Synthetic data generation offers a solution to address this gap; standard pipelines often prompt LLMs using structured user personas, comprising demographics, constraints, and goals, to emulate dialogues. However, current methods relying on coarse demographic attributes often yield generic or stereotyped personas that lack real-world nuance.

To improve the realism of synthetic data, we introduce Socially Grounded Exemplars (SGEs), which translate abstract persona attributes into granular, conversational descriptors. We implemented a two-stage pipeline using GPT-4o to generate SGEs, which then grounded synthetic dialogue generation under various prompting strategies. We evaluated the approach using automatic diversity metrics (Vendi Score) and blinded pairwise preference ratings by community behavioral health specialists (CBHS).

Validation confirmed the feasibility of input generation, with GPT-4o achieving an 85% term acceptability rate for SGEs. In conversation generation, dynamic SGEs significantly improved lexical diversity, achieving a Vendi Score of 289.41 compared to 252.36 for the control baseline. CBHS ranked the model combining dynamic SGEs with implicit name-based cueing highest (Bradley-Terry Score: 0.753), surpassing both the SGE-only model (0.663) and the explicit demographics model (0.348). Raters favored the name-augmented model for “Specificity & Natural Authenticity” (30.0%), while explicit demographic labeling reduced perceived authenticity.

We show SGEs leverage LLM parametric knowledge to produce diverse synthetic data, surpassing the limitations of rigid demographic ontologies. Our findings indicate that implicit cueing through names yields more authentic representations than explicit labeling, reducing the risk of stereotyped outputs. This framework supports the creation of privacy-preserving, conversational datasets informing tasks (e.g. evaluation, agentic workflows, and model distillation) in sensitive healthcare contexts.

## Introduction

Health outcomes are influenced significantly by social, economic, and environmental factors beyond clinical care [1]. Yet, healthcare delivery remains centered on episodic, clinic-based encounters, poorly addressing patients’ ongoing, context-dependent needs [2]. This gap disproportionately affects families facing structural barriers, worsening inequities in health outcomes and access [3]. Health-related social needs (HRSNs), like food insecurity and housing instability, are key indicators of this unmet social risk. Although HRSN screening is growing in clinical settings, connecting identified needs to effective support remains difficult [4]. Families often receive passive information or referrals, forcing them to navigate complex social service systems independently, which limits the real-world impact of screening efforts [5].

Conversational agents, or chatbots, offer a promising, scalable solution to bridge this gap by providing personalized feedback, referral and resource navigation [6–10]. Our prior empirical study with multistakeholder groups and low income households confirmed highly perceived user acceptance but also identified potential barriers to real-world deployment, including considerations about data privacy, inclusivity and personalization, low-resource environments and the infrastructural demands of cloud-based systems [7].

These challenges highlight the need for privacy-preserving models that can operate as personalized assistants in resource-constrained environments. While off-the shelf LLM’s exist, their performance of specialty subdomains, like clinical and HRSN, are reliant on task specific fine-tuning [11]. However, there is a lack of conversational datasets within these subdomains to train and build personalized chatbots to address these needs [9,12]. A viable strategy for addressing these barriers is to use high-quality synthetic conversations, reducing dependence on sensitive user data while supporting development and evaluation of conversational agents [13–15]. Many synthetic dialogue pipelines therefore start from a structured user persona (e.g., demographics, constraints, and goals) and prompt an LLM to generate conversations consistent with that profile [13,16–19]. However, persona fields are often represented as coarse demographic or attribute descriptors (e.g., “homeless,” “benefit recipient”), which are underspecified and can yield generic or caricatured realizations without additional grounding [19–23].

In this paper, we introduce Socially Grounded Exemplars (SGEs), a narrative-based persona augmentation method intended to improve the realism and representational specificity of synthetic conversations for HRSN support. Namely, SGEs are short phrases or descriptions that translate predefined, coarse demographic attributes into specific, conversationally plausible examples. We provide an end-to-end generation procedure that produces SGEs, augments personas, and generates conversations under multiple prompting strategies. We further propose a mixed evaluation design that combines human ratings with automatic validation and diversity analysis to quantify downstream effects. Using this framework, we evaluate whether SGEs are (i) feasible for LLMs to generate at scale, (ii) effective for improving conversation quality and diversity, and (iii) robust when extended from HRSN attributes to nuanced demographic cues such as names [22].

### Relevant Works

Synthetic datasets can be used to support evaluation suites [24,25], robustness stress tests under controlled variations [26–28], and seed conversations for few-shot prompting in low-data settings [29,30]. In this paper, we aim to generate high-fidelity synthetic data to support these diverse applications, including distillation-oriented deployment: transferring capabilities from a large language model to a smaller model that can run locally [31]. Such deployment strategies can mitigate privacy and infrastructure constraints by reducing reliance on cloud-based systems and limiting exposure of sensitive conversational data. However, clinical synthetic data efforts also highlight the need for clearer privacy-risk evaluation and reporting standards, rather than treating synthetic data as automatically “safe” [15].

To be effective in clinically oriented conversational settings, the synthetic corpus must reflect not only task goals but also the interaction patterns that a smaller model should learn, especially how users express HRSNs in everyday language and how they describe constraints, preferences, and follow-up needs [13,16,17]. A common approach is persona-conditioned generation, where prompts include a structured user profile and the model generates dialogue consistent with that profile [13,16,17]. Persona design is often used as a control variable to elicit systematic variation in outputs (e.g., attitudes), which underscores that the form of the persona can materially shape what is generated [18]. In practice, however, personas are frequently built from simple demographic permutations or other coarse attribute labels. This baseline can fail to capture the multidimensionality of identity and can encourage stereotyped or flattened portrayals [20,22]. Moreover, when prompted with explicit demographic labels, LLMs can default to an out-group perspective, and they can struggle with incongruous personas whose attributes do not align with common stereotypes [21,22]. Recent work further shows that expanding personas with model-generated details can introduce skew or homogenization if not carefully designed and evaluated, reinforcing the need for grounded persona representations and explicit quality checks [23].

To improve realism and representational specificity, prior work has enriched user profiles with additional information, such as traits, behavioral attributes, or narrative vignettes [16,17,32]. Related population simulation work similarly argues for methods that better represent heterogeneity across a population rather than producing a single “average” user, including combining multiple persona components to broaden coverage [19]. Building on these directions, we introduce Socially Grounded Exemplars (SGEs) as a targeted augmentation for persona-conditioned dialogue generation: instead of adding more persona fields, we translate abstract persona attributes (e.g., “homeless,” “benefit recipient”) into short, conversationally plausible realizations that can guide generation while remaining tied to the original structured profile. This design is informed by evidence that semantic exemplars can improve control over dialogue generation [33] and that LLM outputs can shift substantially based on small changes in contextual details [34]. By grounding personas in concrete language that could plausibly appear in conversation, SGEs aim to improve specificity, reduce generic outputs, and support more faithful representation in synthetic conversations for HRSN support.

## Methods

Our synthetic conversation pipeline is organized into two generation stages plus a human evaluation stage (Figure 1): Stage 1 (Input Generation) produces socially grounded exemplars (SGEs) and persona inputs for populations with unmet HRSNs, Stage 2 (Utterance Generation + Quality Filtering) produces synthetic conversations and filters them for validity, and CBHS Evaluation compares outputs across methods using a rubric. This structure follows a standard framework for conversational data generation [13]. We target goal-oriented dialogues in which individuals seek community resources from an HRSN-focused chatbot, consistent with prior synthetic dialogue work and prior iterations of HRSN chatbots [7,14]. Across stages, we evaluate outputs for conversational quality and diversity and apply safeguards intended to reduce the risk of harmful bias [20,35].

**Figure 1:**
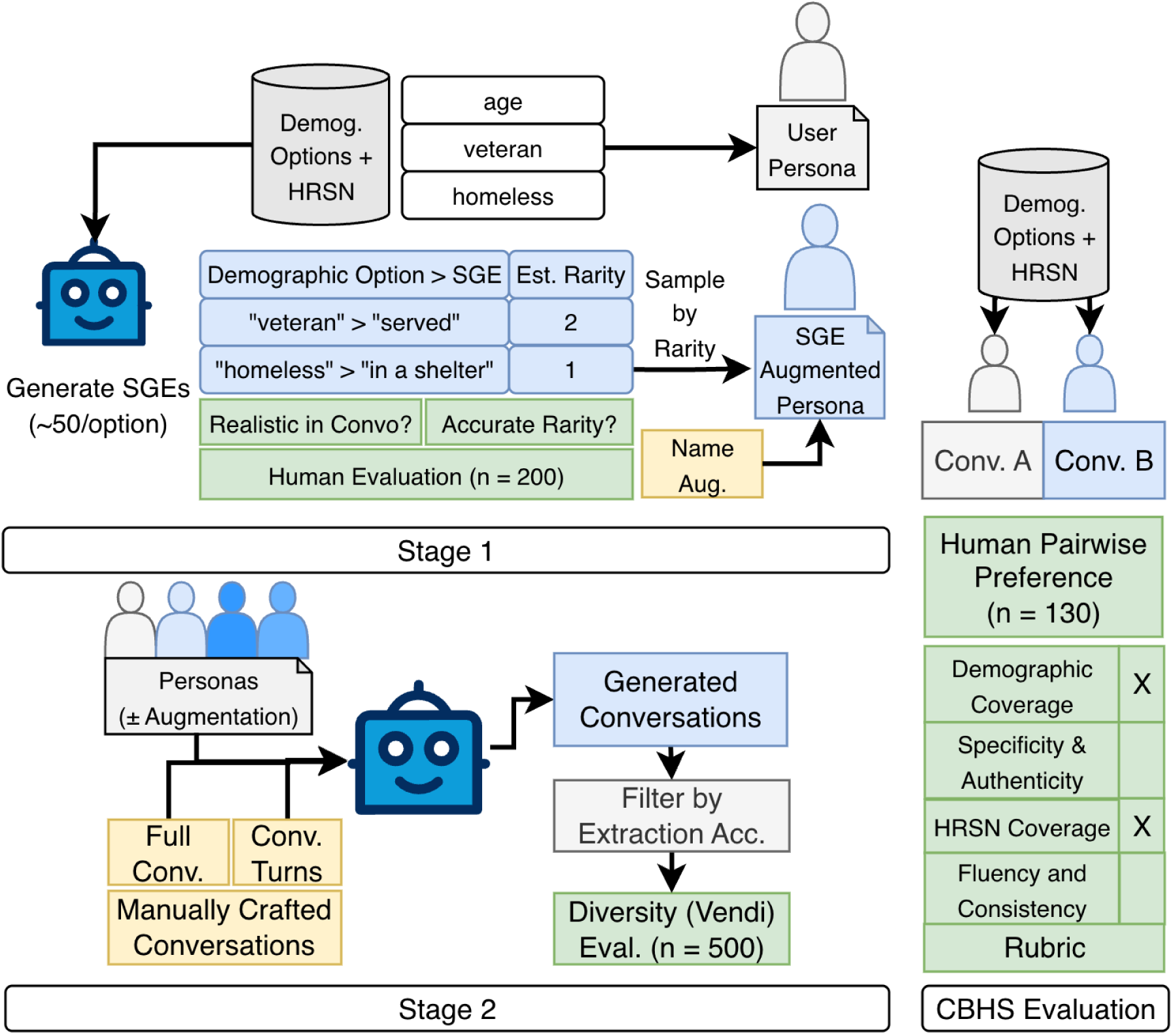
Synthetic data generation pipeline. **Blue**: LLM-generated components; **green**: evaluation; **yellow**: rule-based steps. **Stage 1**: generate SGEs from abstract demographic options, optionally name-augmented, and sample by LLM-assessed rarity. **Stage 2**: use sampled SGEs to construct personas, pair with human-annotated seed conversations (turns or full dialogues), generate synthetic conversations, and filter for extraction accuracy. **CBHS evaluation:** blinded community behavioral health specialist pairwise comparison of conversations (different generation methods given the same seed user profile), with rubric-based justification.

### Stage 1: Socially Grounded Exemplar (SGE) Generation

In this section, we describe the process for creating and validating SGEs. We gather coarse categorical filters from the public-facing social care resource database [36]. The database is designed to help users find a variety of HRSN resources, however we focus on food resources (e.g., community gardens or food pantries). Furthermore, the database can be queried using filters that mark personal status across a set of factors (e.g., housing or income status) to identify relevant nearby resources. A more detailed list of resources, filters, and filter values are shown in Table A1 in the appendix.

For each filter value (e.g., “homeless,” “benefit recipient”), we prompted GPT-4o to generate up to 50 socially grounded exemplars (SGEs): short phrases or descriptions intended to reflect that category and plausibly occur in conversation. This approach uses zero-shot dataset synthesis, where examples are generated directly from a pretrained language model via task-specific prompts rather than collected and annotated from an external corpus, consistent with the general strategy described in ZeroGen [31].

We additionally instructed the model to assign each SGE an estimated rarity score on a 1–5 scale (5 = most rare), intended to approximate how likely the exemplar is to appear in natural dialogue. The full SGE-generation prompt was:

> *“I will provide you a category. These categories are filter options for a website navigating social resources for families in need. Provide a set of common terms that can fit within the category. If relevant, provide terms that focus on pediatric conditions. Provide the common slang for terms if they exist, with a focus on layman phrasing. Provide no more than 50 examples. For each term, quantify how likely the phrasing is in the general population (1–5). Return JSON in the format: {“terms”: [[term_1, rarity], [term_2, rarity], …]}”*

To assess the effect of model capacity on output quality, we performed SGE generation using both GPT-4o and GPT-4o-mini under the same prompt and generation settings. Our evaluation at this stage is based on a subsample of 500 SGEs. Two human annotators independently evaluate each generated SGE for term acceptability (yes/no) based on (i) whether the exemplar fits the intended filter value and (ii) whether it could plausibly occur in informal conversation.

Annotators also independently evaluate rarity-score acceptability (yes/no) by comparing the model-assigned score to their own expectation; a score is marked acceptable if it is within ±1 point on the 1–5 scale. For reporting, we define the term acceptability rate and rarity-score acceptability rate as the proportion of SGEs marked acceptable (per the above criteria), and we compute inter-rater agreement for term acceptability using Cohen’s κ.

### Stage 2: Conversation Generation

In this stage, we create synthetic conversations that emulate conversations between a user seeking resources and an HRSN navigation chatbot.

### User Profile Permutation

We first construct structured user profiles by permuting over the categorical filters, aligning with established “Personalized Profile” methods for synthetic data generation [13,16,17,37]. Each profile was constrained to include 1–4 filter categories, with 1–3 values selected per category, to avoid implausible over-specification that may reflect “incongruous” personas, which LLMs are known to struggle with [21]. Each profile also included a “resource need” sampled from the resource database. The following is an example profile:

> *{resource: [’Help Pay for Food’],*

> *Age Group: [’young adults: 20 - 30 years’],*

> *Family Role: [’DEFAULT’],*

> *Disability: [’limited mobility’, ’visual impairment’],*

> *Income: [’DEFAULT’],*

> *Housing: [‘homeless’],*

> *Health: [’DEFAULT’]}*

Here, ‘*DEFAULT’* emulates cases where no specific filter value is selected. This permutation is provided as an input to emulate a user during conversation generation.

### Human Generated Conversations

We manually authored 50 seed conversations to condition GPT-4o using in-context learning, where a small set of exemplars guides subsequent large-scale synthetic generation [13,38]. Annotators role-played as end users, interacting with an HRSN resource-assistant chatbot (the “Bot”). For each conversation, the annotator was assigned a user profile containing demographic attributes and HRSN factors; profile fields were presented in a randomized order to reduce templated responses. The Bot then presented a scripted sequence of prompts requesting details for specific categories (e.g., “Can you tell me more about your age and housing?”). For each Bot prompt, we specified which profile fields the annotator should incorporate in their reply, and annotators wrote responses in informal, natural language.

Conversations proceeded for 3–4 user turns, after which the protocol ensured that all profile fields had been disclosed. Table 1 provides an example of the annotation schema.

**Table 1:** An example of the annotation schema for manually created conversations. The human annotators are provided the ‘Bot Query’ as well as each element of the user profile to respond with. Elements marked as ‘ignore’ should not be answered in that turn. Elements marked as ‘Don’t answer’ should have the user refuse to answer, either explicitly or by skipping the question. The human annotated responses are in the ‘Human Response’ column

| Bot Query | Resource | Age Group | Disability | Housing | Health | Family Role | Income | Human Response |
| --- | --- | --- | --- | --- | --- | --- | --- | --- |
| Hello! I am here to help you find nutrition resources in your area. How can I help you? | Nutrition Education | ignore | ignore | ignore | ignore | ignore | ignore | I was wondering how I could find out more about what food is good for me |
| Can you tell me more about your age and housing? | ignore | Don't Answer | ignore | runaways | ignore | ignore | ignore | We actually left our parents |
| What about disabilities? | ignore | ignore | visual impairment , physical disability | ignore | ignore | ignore | ignore | My sister doesn't see so well and needs a wheelchair |
| How about considerations regarding health and family role? | ignore | ignore | ignore | ignore | diabetes | dependents , caregiver | ignore | I have diabetes and I take care of my younger sister |

### Conversation Generation Strategy

In the utterance generation stage, we generate 3–6 turn conversations in a single “one-go” call using a shared base prompt [13]. The base prompt sets the interaction format (User–Assistant turns), writing style, and per-turn extraction annotations so that outputs are comparable across conditions. We then vary three prompt components: (1) SGE grounding (none/control, static, dynamic), (2) demographic cueing (none, explicit demographic descriptors, or name-based cues), and (3) few-shot exemplars (full-conversation sampling or turn sampling). Table 2 enumerates the resulting experimental conditions. The following is an example of the base prompt:

> *“You are simulating realistic conversations between a User and a Virtual Assistant supporting nutrition-related resources and programs. Generate a 3–6 turn conversation grounded in the provided Input Profile. User responses should reflect their unique situation and constraints, expressed informally, like text messages, potentially using shorthand or emotion. Embody the user’s perspective, making reasonable inferences based on their Input Profile. After each user response, annotate the ’extracted_values’ dictionary to identify which profile elements were surfaced. Output the entire conversation and annotations as a structured JSON object.”*

**Table 2:** A list of each experiment and the elements modulated.

| Experiment Name | SGE Integration | Demographic Integration | Few-Shot Integration |
| --- | --- | --- | --- |
| Control-Convo | Control | None | Conversation |
| Control-TS | Control | None | Turns |
| Static-Convo | Static | None | Conversation |
| Static-TS | Static | None | Turns |
| Dynamic-Convo | Dynamic | None | Conversation |
| Dynamic-TS | Dynamic | None | Turns |
| Dynamic-Demog-Convo | Dynamic | Demographics | Conversation |
| Dynamic-Demog-TS | Dynamic | Demographics | Turns |
| Dynamic-Names-Convo | Dynamic | Names | Conversation |
| Dynamic-Names-TS | Dynamic | Names | Turns |
| Dynamic-Demog-Names | Dynamic | Both | Conversation |
| Dynamic-Demog-Names-TS | Dynamic | Both | Turns |

### SGE Integration Strategy

We experiment with three distinct strategies for integrating SGEs, which function as semantic exemplars [33] to guide generation: **control**, **static**, and **dynamic**. This approach leverages the known sensitivity of LLMs to specific contextual details, using SGEs as “attractors” to ground the model’s output [34].

The **control** strategy (baseline) omits any explicit exemplar instruction, testing the model’s ability to generate diverse conversations using only the coarse filter categories. The **static** strategy incorporates a fixed exemplar instruction to guide the model’s interpretation of abstract categories. The static instruction aims to isolate whether providing a single SGE is sufficient to ground and improve the diversity of synthetic conversation generation and whether our dynamic approach leads to diminishing returns. An example of the static SGE prompt augmentation is as follows:

> *“[PROFILE VALUE CONCRETIZATION INSTRUCTIONS] When generating User Responses, interpret abstract categories from the Input Profile using specific, realistic examples—e.g., ’Income: benefit recipient’ → ’on SNAP’. These examples help simulate a plausible user situation and make responses more natural. You don’t need to use these examples exactly, but they should guide tone and content. Still annotate ’extracted_values’ based on the original Input Profile keys.”*

The **dynamic** strategy introduces tailored exemplars for every abstract filter category present in the user profile. During prompt generation, the prompt dynamically incorporates specific mappings (e.g., “’Housing: homeless’ → ’living in a shelter’”) based on the generated SGE sets. This strategy is intended to yield richer, more personalized conversations while maintaining variability between conversations.

### Demographic Integration Strategy

We further evaluate the impact of integrating demographic details, applied exclusively with the dynamic SGE condition. The explicit demographics condition assigns detailed descriptors (e.g., “Race: Hispanic”). We sample from these demographics at a rate skewed by the local population distribution (e.g., ‘White’ is sampled at 30% while ‘Asian’ is sampled at 10%). The following is an example demographic prompt augmentation

> *“[USER DEMOGRAPHICS] The user has the following additional background: Race: Hispanic, Gender: Female, Immigrant Status: Yes, Religion: Catholic. These demographics may influence their responses, preferences, and concerns. Reflect this subtly in their language or concerns if appropriate.”*

Conversely, the names condition subtly cues demographics through culturally or gender-associated first names (e.g., “Luis,” “Aaliyah”). This indirect strategy acts as an SGE proxy for demographics, leveraging the model’s pre-trained associations to foster more authentic “in-group” representations and avoid the “flattening” of identity groups [22]. We sample distinctively black and white names by reusing the examples noted in Fyer & Levitt [39] and employed by Wang et al. [22]. Furthermore, we sample the top 5 names for each nationality with large representation among the refugee population of Columbus, OH, as noted by CRIS (e.g. Afghan, Burmese, Somali) [40]. Names for these groups are sampled from the most common of each nationalist, as noted in Forebears [41]. An example name augmentation prompt used was:

> *“[USER NAME] The user’s name is Luis. Use this name to help contextualize the user’s identity and inform your generation of the user’s tone, background, and perspective. The name may suggest cultural, ethnic, or gender associations—use these associations subtly when generating the user’s responses. You do not need to include the name explicitly unless prompted by the Bot.”*

### Few-shot Integration Strategy

Finally, we examine the impact of integrating few-shot exemplars into conversation generation through two distinct sampling methods: full conversation sampling and turn sampling. The full conversation sampling strategy provides GPT-4o with complete exemplar conversations manually created in the “Human Generated Conversations” step, as shown in the example below:

> *“[EXAMPLE CONVERSATION] Input Profile: {’resource’: [’Nutrition Education’], ’Housing’: [’runaways’], ’Disability’: [’visual impairment’]}. Output: [{Bot: ’Can you tell me more about your housing situation?’, User: ’We ran away from home a while ago and stay with friends’, extracted_values: {’Housing’: ’runaways’}}, …]”*

Providing the full conversation illustrates how a user incrementally surfaces their profile details over several turns. The turn sampling strategy presents isolated conversational turns, aiming to promote more flexible and varied dialogue structures. The turn sampling augmentation is constructed as follows:

> *“Below are example TURNS illustrating how users reflect profile values or request clarification.”*

### Conversation Generation Evaluation

Our approach seeks to leverage model associations to diversify generations, but the same mechanism can amplify bias or produce misleading representations. Accordingly, we evaluate both safety-related quality (e.g., realism and representational fidelity) and task-related utility. We design the evaluation to reflect common downstream uses of synthetic dialogue, including evaluation suites, few-shot prompting, and distillation-oriented deployment. In each setting, outputs must be readable and coherent, and they must accurately reflect the input profile so they can support reliable labels and metrics (e.g., extraction accuracy).

### Automatic Evaluation

To gauge the quality and diversity of each conversation generation method, we perform an automatic evaluation of diversity and extraction accuracy. For diversity, we use the Vendi Score to compute lexical diversity (using n-grams, which captures variety in specific word sequences) and semantic diversity (using BERT and simCSE embeddings, which capture variety in underlying meanings) [42]. For extraction accuracy, we apply a factuality check [13], ensuring the extracted values in each turn reflect the input user profile; mismatches are marked as errors and excluded. The top-performing methods are then passed to human evaluation.

### Human Evaluation

Human evaluation occurs in two stages. In the first, annotators perform a pairwise evaluation [13], selecting their preferred conversation from a pair. After selecting a preference, the annotator marks dimensions of quality that justified their decision. These axes, detailed in Table 3, assess fluency and consistency [13] as well as specificity and the authenticity of demographic representation [20,22]. This expert-led evaluation of qualitative factors mirrors approaches used in other specialized, sensitive domains like healthcare [32,38]. We compute inter-rater agreement for conversation preference ratings using Fleiss’ κ.

**Table 3:**
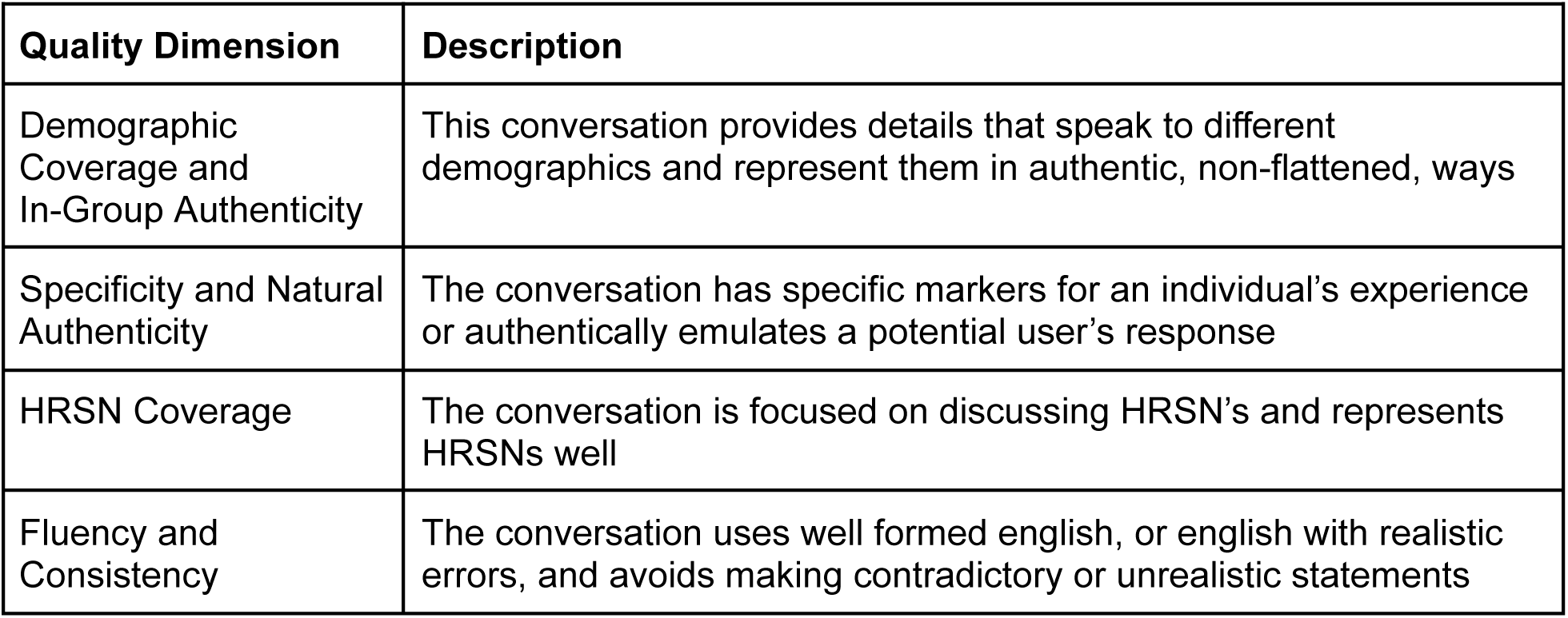
Dimension of Quality for human evaluation.

## Results

**Stage 1** evaluated the feasibility of generating SGEs using LLMs. In human review of the pre-specified set of 500 SGEs per model, GPT-4o achieved a term acceptability rate of 85%, while GPT-4o-mini scored a substantially lower 69%. GPT-4o’s rarity scores were acceptable 95% of the time under the ±1-point criterion, compared to 92% for GPT-4o-mini. Inter-rater agreement for term acceptability was substantial (κ = 0.75). These results indicate that SGEs can be produced efficiently via prompting, however output quality is sensitive to model capacity.

In **Stage 2**, we evaluated the synthetic conversations generated using the SGEs. This evaluation was performed in two phases: an automatic assessment of conversational diversity and a human-powered evaluation of conversational quality and preference. To automatically assess conversational diversity, we used the Vendi Score, a metric that measures the lexical (n_gram) and semantic (BERT, simCSE) variety in a set of generated texts. As shown in Table 4, the prompting strategy had a clear impact on diversity (higher scores denote greater diversity). Using turn-sampling as a few-shot integration strategy consistently produced greater lexical diversity than using full-conversation sampling. In aggregate, we see that turn sampling improved lexical diversity by an average of 11.89% (1.31% STD).

**Table 4:** Vendi calculated diversity metrics (n_gram, BERT, and simCSE) reported for each experimental group on 500 conversations. TS refers to turn-sampling. Higher scores note greater diversity.

| experiment_name | SGE | Demog | Sampling | n_gram | BERT | simCSE |
| --- | --- | --- | --- | --- | --- | --- |
| control | control | none | convoSampling | 227.99 | 1.33 | 35.63 |
| control-TS | control | none | turnSampling | 252.36 | 1.33 | 35.85 |
| static | static | none | convoSampling | 241.68 | 1.31 | 35.43 |
| static-TS | static | none | turnSampling | 264.96 | 1.32 | 34.26 |
| dynamic | dynamic | none | convoSampling | 252.53 | 1.33 | 36.15 |
| dynamic-demog | dynamic | demog | convoSampling | 264.72 | 1.32 | 35.55 |
| dynamic-demog-name | dynamic | demog | convoSampling | 254.20 | 1.33 | 35.05 |
| dynamic-demog-name-TS | dynamic | both | turnSampling | 287.84 | 1.33 | 32.37 |
| dynamic-demog-TS | dynamic | demog | turnSampling | 296.07 | 1.31 | 33.28 |
| dynamic-name | dynamic | name | convoSampling | 251.29 | 1.32 | 34.28 |
| dynamic-name-TS | dynamic | name | turnSampling | 283.70 | 1.32 | 33.18 |
| dynamic-TS | dynamic | none | turnSampling | 289.41 | 1.33 | 35.13 |

The SGE integration strategy also improved lexical diversity. The static-TS model (264.96, 4.87% above control) was more diverse than the control-TS baseline (252.36), and the dynamic-TS model (289.41, 13.68% above control) showed an even greater increase. Notably, the effect of SGEs was strong enough that the dynamic-convo model (252.53), which did not use turn-sampling, achieved a lexical diversity score nearly identical to the control-TS baseline (252.36), indicating that the SGEs themselves provide a significant diversity enhancement. The dynamic-demog-TS model, which combined dynamic SGEs with explicit demographic data, yielded the highest lexical diversity score (296.07). Across all experiments, semantic diversity scores remained stable (e.g., BERT Vendi scores ranged from 1.31 to 1.33), suggesting that while the wording of the conversations became more varied, the core topics appropriately remained focused on HRSN navigation.

Following the automatic evaluation, we conducted a human preference evaluation on the four most lexically diverse models, all of which used turn-sampling. Four social worker raters performed a blind, pairwise comparison of 130 conversations. We calculated a Fleiss’ Kappa of 0.491, indicating moderate agreement. This level of consensus is typical for subjective evaluations of open-ended dialogue and supports the reliability of the resulting preference trends [43,44]. We analyzed the results using a Bradley-Terry (BT) ranking model, which calculates a preference score for each model based on how often it was selected as “better” than its competitors. Table 5 displays these preference rankings. The Dynamic-Turn + Name model, which used SGEs and implicit demographic cues (names), was the clear winner, achieving the highest preference Rank 1 (BT Score: 0.753). The Control-Turn model, which used no SGEs or demographic data, was ranked last (Rank 4) and served as the baseline (BT Score: 0). The Dynamic-Turn model, using SGEs alone, placed second (Rank 2, BT Score: 0.663). Critically, the Dynamic-Turn + Demog model, which used explicit demographic labels (e.g., “Race: Hispanic”), was ranked third (BT Score: 0.348), performing substantially worse than the name-based or SGE-only models.

**Table 5:** Human preference scores through pairwise comparison of the most diverse systems, reported through Bradley-Terry Ranking. Scores were assessed by four social worker raters in a blind evaluation of 130 conversations. Fleiss’ Kappa (κ): 0.491

| Context Type | Mean Rank | Mean BT Score | Confidence Interval |
| --- | --- | --- | --- |
| Control-Turn | 4 | 0.000 | [0.000 - 0.000] |
| Dynamic-Turn | 2 | 0.663 | [0.349 - 0.977] |
| Dynamic-Turn + Demog | 3 | 0.348 | [0.037 - 0.659] |
| Dynamic-Turn + Name | 1 | 0.753 | [0.441 - 1.065] |

Raters additionally noted the rationale for their preference, as outlined in Table 6. Raters selected the winning Dynamic-Turn + Name model most frequently for ’Demographic Coverage and In-Group Authenticity’ (34.8%) and ’Specificity & Natural Authenticity’ (30.0%). This model, along with the Dynamic-Turn model, also scored highest on ’HRSN Coverage’ (34.1% and 35.2%, respectively). The Control-Turn baseline scored lowest across all categories, while the explicit Dynamic-Turn + Demog model was notably deficient in ’HRSN Coverage’ (15.6%) and ’Demographic Coverage’ (22.4%), suggesting raters found these conversations less authentic and less relevant.

**Table 6:**
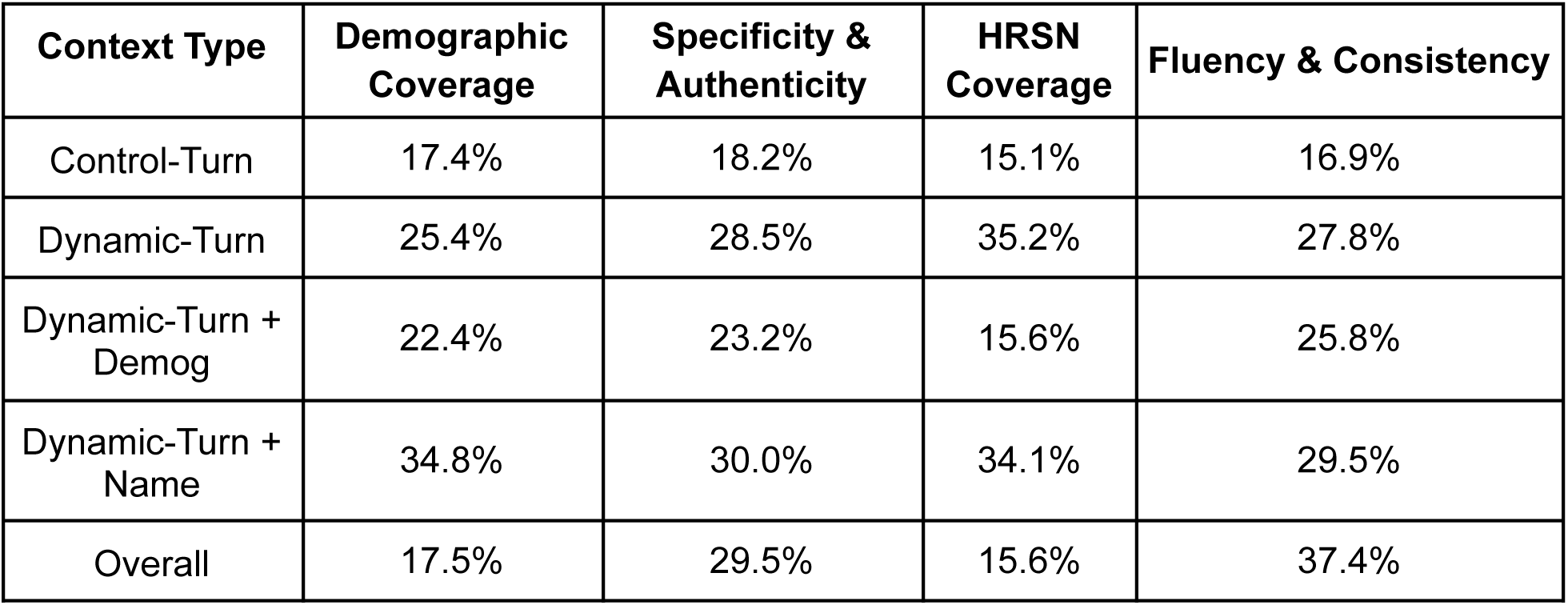
Reasons listed for human-evaluated pairwise preference of conversations. Reported as a percentage of conversations averaged over the four raters (n=130)

## Discussion

This study introduced and evaluated SGEs as a method to improve the quality and representativeness of synthetic conversational data for HRSN-oriented chatbots. Our primary motivation is supporting broad conversational model development, where the resulting data and pipeline facilitate diverse workflows such as few-shot seeding, model distillation, and robustness testing. Our findings show that SGEs are feasible to generate at scale using LLMs and that they improve the realism and diversity of synthetic conversations, including for nuanced demographic representations. Collectively, our main contributions are the introduction and validation of SGEs as an effective method for enriching synthetic personas, the demonstration of their positive impact on conversation realism and diversity, and the crucial finding that implicit demographic cueing through names outperforms explicit labeling for generating authentic representations.

Human validation confirmed the feasibility of generating high-quality SGEs via LLMs, byGPT-4o achieving an 85% acceptability rate for generated exemplars. This result establishes a scalable method for the crucial “input generation” stage within standard synthetic data pipelines [13], providing the kind of rich, contextualized input needed to generate high-fidelity simulations for tasks ranging from rigorous evaluation to knowledge distillation [24,25,31]. While feasible, the observed difference in quality between GPT-4o and the smaller GPT-4o-mini suggests that leveraging more capable foundation models is beneficial for generating the initial SGEs.

We examined the impact of SGEs and different prompting strategies on conversation diversity and quality. Both automatic metrics and human evaluations showed benefits from SGE integration in terms of diversity, consistency, and authenticity. Lexical diversity, measured by n-gram Vendi scores, increased notably with the use of SGEs, especially dynamic ones, and was further enhanced by using turn-sampling (TS) instead of full-conversation sampling as few-shot examples (Table 4). The consistent gains from turn-sampling suggest that providing isolated examples of interaction encourages more varied phrasing and conversational structures compared to constraining the model with entire dialogue trajectories. Semantic diversity remained stable across conditions, indicating that the increased lexical variety stayed appropriately focused on the core HRSN navigation task.

Human preference evaluations strongly corroborated these findings. Expert raters (CBHS) overwhelmingly preferred conversations generated with dynamic SGEs over the control baseline (Table 5). This preference was primarily attributed to enhanced “Specificity & Natural Authenticity” and “HRSN Coverage” (Table 6), suggesting SGEs help LLMs move beyond generic responses and generate dialogues that feel more personalized and relevant to the user’s specific social needs. This directly addresses the known issue of LLMs producing flattened, stereotypical “caricatures” when simulating personas [20]. Our use of CBHS with care coordination responsibilities as raters, given their domain expertise and sensitivity to social context, may also provide a more nuanced evaluation than standard crowdsourcing, potentially identifying subtle biases that less informed raters might miss [35].

We explored how the SGE approach could capture nuanced demographic identities. The findings provide a strong endorsement for using implicit cues over explicit labels. The model using identity-coded names (Dynamic-Turn + Name) was ranked highest by expert raters (Rank 1, Table 5). The Dynamic-Turn + Demog model, using explicit labels, ranked significantly lower (Rank 3). Rationale analysis (Table 6) showed the name-based model excelled in ’Demographic Coverage and In-Group Authenticity’. This result provides compelling evidence in an open-ended generative task that supports the hypothesis of Wang et al. [22]: explicit demographic prompts risk triggering biased, “out-group” representations. In contrast, the identity-coded names act as SGE proxies for demographics, implicitly cueing the model through its pre-trained associations. In other words, in the data models are pretrained on, demographic labels are often used when speaking of a group, while names are often used by the speaker themselves [22]. Leveraging these implicit relationships fosters more authentic, “in-group” portrayals, avoiding the pitfalls associated with prompting LLMs to embody “incongruous” personas [21] or triggering harmful stereotypes [20]. By pairing dynamic SGEs for social context with these name proxies for identity, our method leverages the LLM’s parametric knowledge in a nuanced way, steering generation towards more authentic interactions.

The SGE-augmented synthetic data generated through our pipeline shows promise diversity and authenticity robustness in conversational model development, an area which actively leverages emulated data [24,25,28–31]. While purely synthetic data can offer advantages in diversity and coverage, particularly for rare scenarios, the most robust approach for deploying chatbots in complex, sensitive domains like healthcare may ultimately involve hybrid datasets that combine the breadth of synthetic data with the specific grounding only real user interactions can provide [38].

### Limitations and Future Work

Our study is subject to limitations. Results are based on a single LLM family (GPT-4o) and a specific HRSN domain. The evaluation relied on simulated conversations and a small pool of expert raters, and primarily focused on the user simulation aspect. While SGEs harness the power of parametric knowledge efficiently, this reliance also requires careful validation to ensure that underlying model biases are mitigated rather than amplified. While our results suggest that dynamic SGE generation combined with implicit naming and human oversight offers a promising path towards this goal, we do not currently employ bias validation beyond human expert ratings. Finally, we have not employed automated bias evaluation methods and compared between LLM models in terms of bias propagation.

Future work should continue efforts towards bias mitigation through stakeholder and expert evaluation of conversations and downstream models. Additionally, human annotation can be paired with recent advances in bias detection, such as using semantic probes to ensure no subgroup is considered in more negative terms than another [37,45]. At all steps, it will be important to consider LLMs which are validated to have less stereotyped representations [46]. In addition, future work should investigate the efficacy of these synthetic conversations in downstream applications (e.g. prompt augmentation, distillation, and automatic evaluation). Exploring agentic self-play frameworks [14,32] and integrating richer narrative vignettes [32] could further enhance realism. Grounding generation through API integration with live resource databases and conducting real-world user testing with a distilled chatbot are crucial next steps towards deploying equitable and effective conversational tools for vulnerable populations.

## Conclusion

This work introduces and validates Socially Grounded Exemplars as a novel, LLM-driven method for generating higher-quality synthetic conversational data for HRSN resource navigation chatbots. Our findings demonstrate that SGEs are feasible to generate automatically and that integrating them dynamically improves lexical diversity and results in conversations perceived by domain experts as more authentic and relevant compared to baseline methods lacking specific contextual grounding. Furthermore, we show that using implicit demographic cues via identity-coded names serves as an effective SGE proxy, yielding more authentic “in-group” representations preferred over explicit demographic labeling, extending findings from prior work on bias and persona representation [21,22]. These contributions offer a valuable approach for researchers and practitioners seeking to create diverse, realistic synthetic datasets to support the development and evaluation of equitable, privacy-preserving conversational agents, particularly for deployment in sensitive, resource-constrained clinical and community settings.

## Data Availability

All synthetic conversations and SGEs used for experimentation are available on GitHub (https://github.com/Amad881/SGE_Synth_Convos).

## Appendix

**Table A1:** Example HRSN services and filter options used to navigate our HRSN database. We use these values to inform our user profiles prior to SGE generation.

| Services |  | Filter | Filter Value Examples |
| --- | --- | --- | --- |
| Community Garden |  | Age Group | Infants, toddlers, preschoolers, seniors |
| Food Delivery |  | Disability | Developmental, limited mobility, all disabilities |
| Food Pantry |  | Housing | Homeless, near homeless, runaways |
| Help Pay for Food |  | Health | Alzheimer's, cancer, chronic illness, seizure |
| Meals |  | Role | Parents, caregivers, spouses, dependents |
| Nutrition Education |  | Income | Benefit recipient, low-income |

